# Socio-demographic and environmental factors amplify typhoon-related excess mortality in Japan

**DOI:** 10.64898/2026.09.01.26362002

**Authors:** Lisa Yamasaki, Hiroaki Murayama, Paul LC Chua, Masahiro Hashizume, Robbie M. Parks

## Abstract

Mechanisms shaping population vulnerability to typhoon-related mortality remain poorly understood. Constructing a Bayesian spatio-temporal model, we linked 12.9 million deaths across Japan from 2010 to 2019 to population-weighted typhoon wind exposure and assessed effect modification by income, natural hazard vulnerability and healthcare access. Typhoon exposure was associated with 2,426 cumulative excess deaths [95% credible interval (CrI): 139, 4,632] among adults ≥70 years, with mortality increasing within 0-1 weeks of exposure and more strongly in areas with limited healthcare access and greater hazard vulnerability. Among individuals <70 years, cumulative excess mortality was uncertain [781 deaths; −309 to 1,900], but delayed mortality increases were concentrated in lower-income and landslide-prone areas. These distinct patterns suggest that typhoon mortality reflects an interaction between acute exposure, demographic ageing and geographically uneven adaptive capacity, highlighting the need to incorporate local vulnerability into climate-resilient health systems.

## Main

Typhoons and other tropical cyclones have a devastating impact on mortality in many areas across the world.^1–4^ Globally, 430 million people worldwide, about 5.5% of the world population, were exposed to tropical cyclones in 2024,^5^ causing extensive damage to lives and infrastructures.^6^ With climate projections indicating that tropical cyclones are likely to become more intense as ocean temperatures rise,^7,8^ understanding of the underlying mechanisms through which these events give rise to new deaths is crucial for developing optimal prevention and mitigation strategies.

Tropical cyclone–related mortality arises through complex direct and indirect pathways that vary across geographic, demographic, infrastructural and healthcare contexts. Direct causes such as storm surges, flooding and structural collapse typically dominate mortality in low- and middle-income countries, where inadequate housing and limited healthcare access place vulnerable groups such as infants and young children at particular risk.^9^ In high-income countries, while direct trauma deaths from typhoons still occur,^2^ resilient infrastructure and early warning systems have substantially mitigated their impact, making indirect and delayed causes, such as evacuation-related stress and exacerbation of pre-existing chronic conditions,^10^ a more prominent component of the overall mortality burden. Mortality is often higher in low- and middle-income countries due to inadequate housing and limited healthcare access for vulnerable groups such as infants and young children, in contrast to high-income countries where these indirect pathways may be especially important amid ageing populations and high chronic disease prevalence.

Japan, the world’s most ageing nation, with 29.4% of its population aged 65 years or older in 2025, and a country with one of the highest exposures to tropical cyclones, represents a frontline where demographic vulnerability intersects with climate hazards. Older adults are particularly vulnerable to climate disasters.^11,12^ However, mortality risks among older populations may differ markedly between urban and rural settings due to variations in preparedness and social support,^13^ geographic factors such as flood- or landslide-prone terrain,^14^ housing characteristics and access to medical care, which underscores the need to identify key environmental and social determinants and helps guide targeted disaster risk-reduction and continuity-of-care strategies.

Here, we quantified the age-specific relative contributions of multiple factors associated with tropical cyclones in Japan, a country of 126 million people as of 2019,^15^ with one of the world’s highest exposures to typhoons,^6^ an extremely ageing society and distinct geographical vulnerabilities such as some densely populated flood- and landslide-prone areas.

## Results

### Typhoon exposure

During 2010 to 2019, 579 weeks of typhoon exposure were identified across all 47 prefectures. Annually, there were a range of 9 to 117 prefecture-weeks of exposure with a mean of 58.6 each year. Typhoons were most frequent in Okinawa—Japan’s southernmost prefecture located in the primary typhoon-forming region near the Philippine Sea—with 29 total weeks of exposure, followed by other southern coastal prefectures (Figure 1A). All typhoon exposure occurred during May to October, with a peak in August (154 prefecture-weeks; 26.7% of the total during the study period) coinciding with the warmest season in Japan (Figure 1B, C).

**Figure 1.**
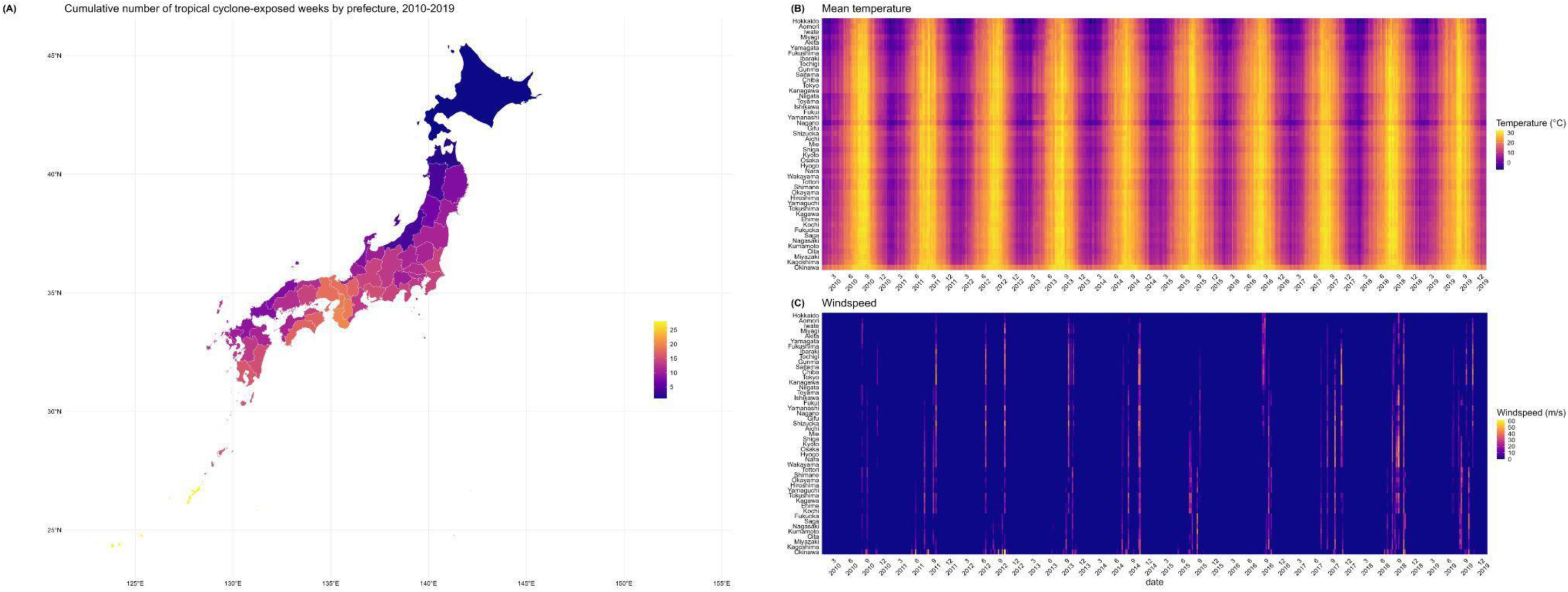
Spatial and temporal distribution of typhoon exposure and meteorological conditions in Japan, 2010–2019. (A) Cumulative number of typhoon-exposed weeks by prefecture during the study period (2010-2019) (B) Daily mean temperature and (C) daily sustained maximum wind speed by prefecture

### Study population and socioeconomic context

During 2010–2019, Japan recorded a total of 12,932,272 deaths, of which 2,363,786 (18.3%) occurred among those aged <70 years and 10,568,486 (81.7%) among those aged ≥70 years (Table 1).

**Table 1.** Typhoons affecting Japan during 2010–2019: maximum wind speeds and total deaths in the younger and older populations during the typhoon week each year.

| Year | Typhoons (maximum windspeed, m/s) | Typhoon exposures (prefecture-week) | Total deaths among those under 70 years | Total deaths among those over 70 years |
| --- | --- | --- | --- | --- |
| 2010 | Dianmu (21.7), Malou (31.9) | 9 | 409 | 1694 |
| 2011 | Aere (21.8), Ma-on (34.7), Muifa (43.2), Roke (49.9), Songda (44.2), Talas (27.0) | 53 | 5451 | 19189 |
| 2012 | Bolaven (42.3), Guchol (40.6), Haikui (20.1), Jelawat (61.7) | 53 | 6590 | 23714 |
| 2013 | Danas (32.2), Francisco (17.2), Man-yi (44.8), Toraji (29.0), Wipha (26.9) | 49 | 5298 | 21293 |
| 2014 | Halong (38.3), Nakri (27.8), Neoguri (28.3), Vongfong (42.5) | 86 | 8021 | 33845 |
| 2015 | Etau (24.5), Goni (49.4), Halola (29.5) | 33 | 1776 | 8560 |
| 2016 | Chaba (28.1), Malakas (35.2), Mindulle (32.2), Namtheun (32.3) | 43 | 4046 | 18335 |
| 2017 | Lan (52.3), Nanmadol (28.4), Noru (28.8), Saola (30.4), Talim (44.7) | 75 | 6398 | 32236 |
| 2018 | Gaemi (25.9), Jebi (50.2), Jongdari (33.4), Leepi (20.0), Maria (18.5), Prapiroon (25.0), Rumbia (32.5), Trami (50.4) | 117 | 9598 | 48676 |
| 2019 | Faxai (34.5), Francisco (26.4), Hagibis (46.9), Krosa (35.0), Nari (18.6), Tapah (36.1) | 61 | 5437 | 29347 |
| Total |  | 579 | 53024 | 236889 |

Income was highest in Tokyo and low in southern and northern prefectures (Supplementary Figure 1A–D). Potential flood risk was generally high in the central main island, with the highest value observed in Tokushima prefecture in Shikoku. Potential landslide risk was elevated in southwestern prefectures, including Hiroshima and Kochi. Access to medical facilities was highest in major metropolitan areas, particularly around Tokyo and Osaka.

### Nationwide excess mortality

Over the 10-year study period (2010–2019), the estimated nationwide cumulative excess deaths were 2,426 [95% CrI: 139, 4,632] among adults aged ≥70 years and 781 [95% CrI: −309, 1,900] among individuals aged <70 years. The posterior probabilities that cumulative excess deaths exceeded zero were 98.0% and 92.0%. In contrast, national-level excess mortality associated with individual typhoons was highly uncertain (Supplementary Table 3). Among adults ≥70 years, the largest posterior medians were148 [95% CrI: −116, 449] for Typhoon Vongfong in 2014, followed by Typhoon Lan in 2017 [91; −85, 275], Typhoon Hagibis in 2019 [82; −36, 203] and Typhoon Roke in 2011 [52; −74, 187]. Among people <70 years, posterior medians for the same events were 55 [95% CrI: −93, 224], 49 [95% CrI: −33, 135], 39 [95% CrI: −11, 92] and 50 [95% CrI: −32, 139]. All 95% credible intervals included zero with substantial uncertainty in single-event national aggregation.

### Typhoon-prefecture excess deaths

Among those aged <70 years, excess deaths were distributed across multiple prefectures and typhoons, with consistent signals—defined as posterior probability >95% across multiple events—in Okinawa, Ibaraki, and Chiba (Figure 2A). Among those aged ≥70 years, excess deaths were concentrated in a few prefectures with substantially larger absolute magnitudes, particularly Chiba, Tokyo, and Kanagawa. The large signals were observed during Vongfong (2014), Lan (2017), and Roke (2011); the absolute magnitude was also markedly larger than in the younger group (Figure 2B).

**Figure 2.**
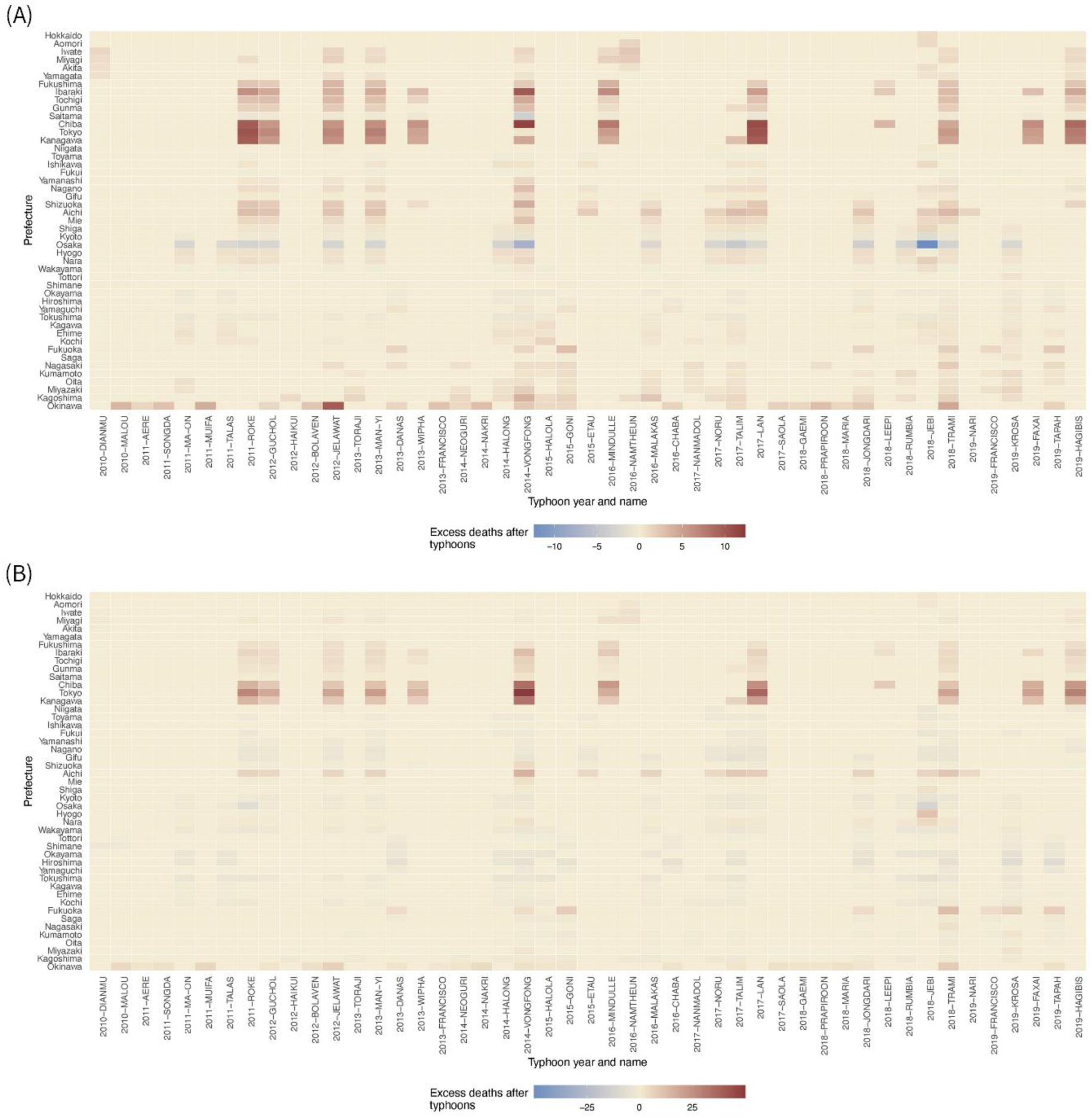
Prefecture-specific excess deaths by typhoon name for younger (<70 years) and older (≥70 years) population. (A) the younger population aged <70 years and (B) the older population aged ≥70 years.

Among <70-year-olds, Ibaraki had excess deaths across multiple typhoons—Roke (2011) at 6 [95% credible interval (CrI) 1, 11], Guchol (2012) at 4 [95% CrI: 1, 8], Vongfong (2014) at 10 [95% CrI: 2, 18], Mindulle (2016) at 7 [95% CrI: 2, 11], and Lan (2017) at 5 [95% CrI: 1, 9] (all >99% probability) (Supplementary Table 1). Okinawa showed similarly consistent signals, with 10 [95% CrI: 2, 18] (>99%) during Jelawat (2012), reflecting its 29 typhoon-weeks of exposure. Excess mortality also extended to the inland prefectures of Tochigi (cumulative 57 [95% CrI: 19, 90]; 100% probability) and Gunma (37 [95% CrI: 9, 63]; 99.8%). The largest estimates were Chiba during Vongfong (2014) at 12 [95% CrI: −5, 29] and during Lan (2017) at 11 [1, 22] (Table 2).

**Table 2.**
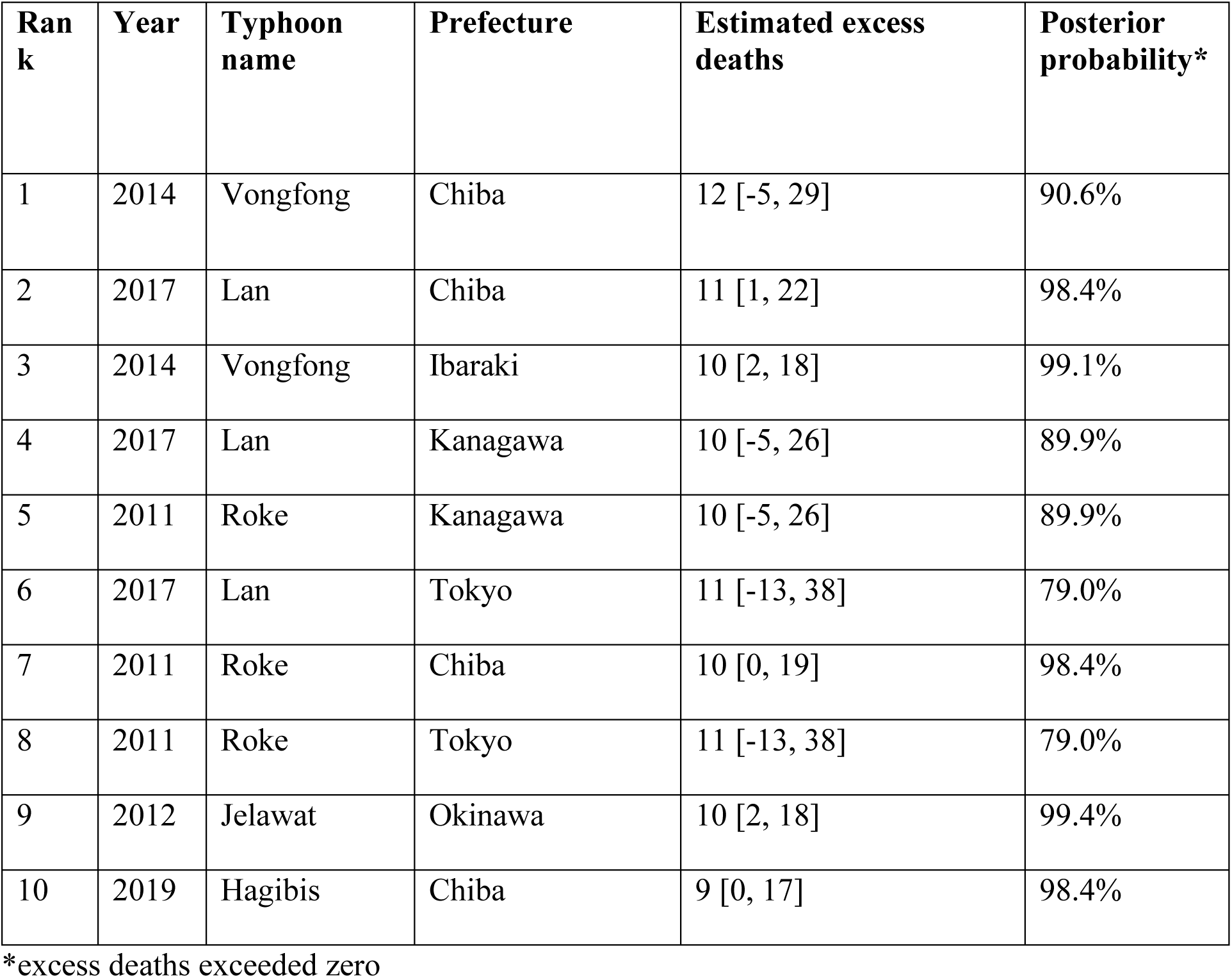
Estimated typhoon-& prefecture-specific excess deaths after the top 10 most deadly typhoons in the group aged under 70 years. *excess deaths exceeded zero.

Among ≥70-year-olds (Table 3, Figure 2B), Chiba during Vongfong (2014) at 35 [95% CrI: 6, 64] (>95% probability) and Tochigi at 7 [95% CrI: 1, 13] (>95% probability) had 95% CrI excluding zero. Chiba also showed consistent signals across multiple typhoons, including Lan (2017) at 25 [95% CrI: 4, 46], Hagibis (2019) at 23 [95% CrI: 3, 42], and Mindulle (2016) at 23 [95% CrI: 4, 41], all >99% probability (Supplementary Table 2). Okinawa showed consistent signals across multiple typhoons (Figure 2B; Supplementary Table 2). Negative point estimates were observed in Osaka across both age groups (Figure 2A, B).

**Table 3.** Estimated typhoon- & prefecture-specific excess deaths after the top 10 most deadly typhoons in the group aged over 70 years. *excess deaths exceeded zero.

| **Ran**<br>**k** | **Year** | **Typhoon**<br>**name** | **Prefecture** | **Estimated excess**<br>**deaths** | **Posterior**<br>**probability\*** |
| --- | --- | --- | --- | --- | --- |
| 1 | 2014 | Vongfong | Tokyo | 50 [-27, 135] | 90.0% |
| 2 | 2017 | Lan | Tokyo | 37 [-14, 94] | 90.2% |
| 3 | 2014 | Vongfong | Chiba | 35 [6, 64] | 98.8% |
| 4 | 2014 | Vongfong | Kanagawa | 33 [-11, 81] | 92.4% |
| 5 | 2019 | Hagibis | Tokyo | 29 [-11, 75] | 90.2% |
| 6 | 2011 | Roke | Tokyo | 27 [-11, 71] | 90.2% |
| 7 | 2017 | Lan | Chiba | 25 [4, 46] | 99.1% |
| 8 | 2016 | Mindulle | Chiba | 23 [4, 41] | 99.0% |
| 9 | 2019 | Hagibis | Chiba | 23 [3, 42] | 99.1% |
| 10 | 2013 | Man-yi | Tokyo | 22 [-8, 55] | 90.2% |

\*excess deaths exceeded zero

### Lag-specific effect modification by socio-economic and geographic factors on excess mortality among younger people

The posterior probability that cumulative excess deaths exceeded zero among those aged <70 years was high (>0.8) across eastern Japan, with the exception of the greater Tokyo area, but was substantially lower across western Japan (Supplementary Figure 2A). During 2010-2019, cumulative excess deaths among the younger population were geographically distributed, with particularly high totals observed in southern and eastern regions along the Pacific coast. Ibaraki experienced the largest cumulative excess deaths at 133 [95% CrI: 56, 206] deaths (Figure 3A). On a population-standardised basis, the cumulative excess death rate was concentrated in a relatively narrower set of predominantly coastal prefectures. Northeastern prefectures near Tokyo showed elevated burdens, including Ibaraki (6 per 100,000 [95% CrI: 2, 9]) and Fukushima (4 per 100,000 [95% CrI: 2, 6]). Similarly, higher population-adjusted death rates were observed along the Pacific-facing southern coast, including Kochi (4 per 100,000 [95% CrI: −1, 9]), Miyazaki (4 per 100,000 [95% CrI: 2, 7]), Kagoshima (6 per 100,000 [95% CrI: 1, 10]), and Okinawa (9 per 100,000 [95% CrI: 1, 16]) (Figure 3B). Okinawa had the highest population-standardised excess death rate among those aged <70 years (9 per 100,000 [95% CrI: 1, 16]; 98.9% probability), despite also showing an elevated rate among ≥70 years (46 per 100,000 [95% CrI: −20, 105]; 92.6% probability), underscoring compounding vulnerability across all age groups in this island prefecture exposed to 29 typhoon-weeks over the study period. Prefecture-level estimates of the overall death rate change per 1 m/s increase in windspeed varied geographically (Figure 3C). For example, increases were observed in some southern parts of Japan including Kagoshima (0.27% [95% CrI: 0.07, 0.48]) and Okinawa (0.22% [95% CrI: 0.02, 0.41]), as well as Ibaraki (0.27% [95% CrI: 0.12, 0.41]), located in the eastern Pacific coast. Some northern prefectures also exhibited higher wind-related vulnerability with mortality rate increase of 0.25% [95% CrI: 0.12, 0.38] in Aomori, 0.30% [95% CrI: 0.16, 0.44] in Iwate and 0.13% [95% CrI: 0.03, 0.25] in Miyagi, although the overall variation in mortality remained modest.

**Figure 3.**
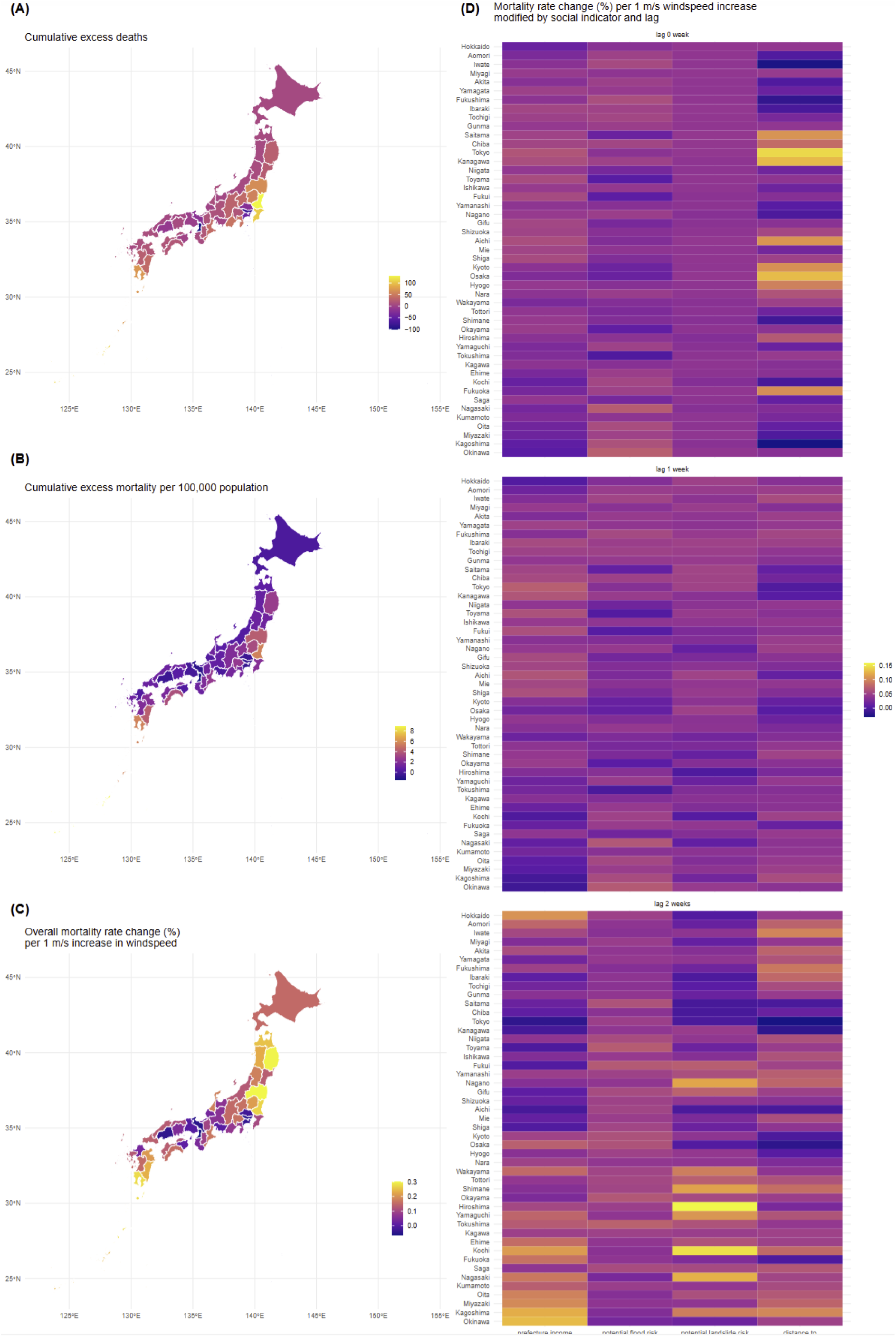
Typhoon-related mortality burden and its modification by socio-environmental factors in the population aged <70 years. (A) Cumulative excess deaths by prefecture, 2010– 2019. (B) Cumulative excess mortality rate (%). (C) Overall mortality rate change (%) per 1 m/s increase in windspeed. (D) Modification of mortality risk (percent change per 1 m/s windspeed increase) by prefecture-level vulnerability indicators (income, flood risk, landslide risk, access to medical facilities) at 0, 1 and 2-week lags

Among people <70 years, modifier effects on excess mortality varied across post-typhoon weeks, with the most geographically extensive associations emerging at the two-week lag. In the week of a typhoon, clear associations were observed for distance to medical facilities, with higher mortality in major metropolitan areas: 0.14% [95% CrI: 0.05, 0.24] in Tokyo and 0.13% [95% CrI: 0.04, 0.21] in Osaka for younger generations (Figure 3D). At 2 weeks after typhoons, associations were strongest for potential landslide risk, with higher mortality observed in Hiroshima (0.16% [95% CrI: 0.04, 0.28]), Kochi (0.15% [95% CrI: 0.04, 0.26]), and Nagasaki (0.12% [95% CrI: 0.03, 0.21]). Income level also modified delayed mortality risks: in southern Japan, economically disadvantaged southern areas such as Kagoshima and Okinawa experienced higher mortality at lag 2 weeks, with mortality increases of 0.12% [95% CrI: 0.04, 0.20] and 0.13% [95% CrI: 0.04, 0.21], respectively. In contrast, prefectures characterised by potential flood risk showed minimal or no increase.

### Lag-specific effect modification by socio-economic and geographic factors on excess mortality among older people

Among those aged ≥70 years, high posterior probabilities (>0.8) were more spatially concentrated than among the younger group, particularly in northeastern Honshu and Kanto prefectures including Iwate, Miyagi, Fukushima, Ibaraki, Tochigi, Gunma, Chiba, Tokyo, and Kanagawa (Supplementary Figure 2B). Excess deaths among the older population were largest by point estimate in Tokyo (464 [95% CrI: −219, 1143]; 90.1% probability), with Chiba showing the highest cumulative burden clear of null at 366 [95% CrI: 85, 659] (99.3% probability) (Figure 4A). The inland prefectures of Tochigi and Gunma also ranked high nationally, with cumulative excess deaths of 101 [95% CrI: 32, 163] (99.8% probability) and 71 [95% CrI: 13, 125] (99.4% probability), respectively (Figure 4A). The death rate showed a pattern similar to that observed in the younger population, with higher rates in coastal prefectures northeast of Tokyo (e.g. 39 per 100,000 [95% CrI: 12, 65] in Ibaraki and 33 per 100,000 [95% CrI: 8, 60] in Chiba), and a comparable rate of 46 per 100,000 [95% CrI: −20, 105] in Okinawa (Figure 4B). When wind speed intensity increased, the most vulnerable regions in Japan largely overlapped with those showing high excess mortality, particularly along the northeastern Pacific coast, as shown in Figure 4C. The inland prefectures of Tochigi and Gunma also showed elevated wind-speed-related vulnerability (Figure 4C).

**Figure 4.**
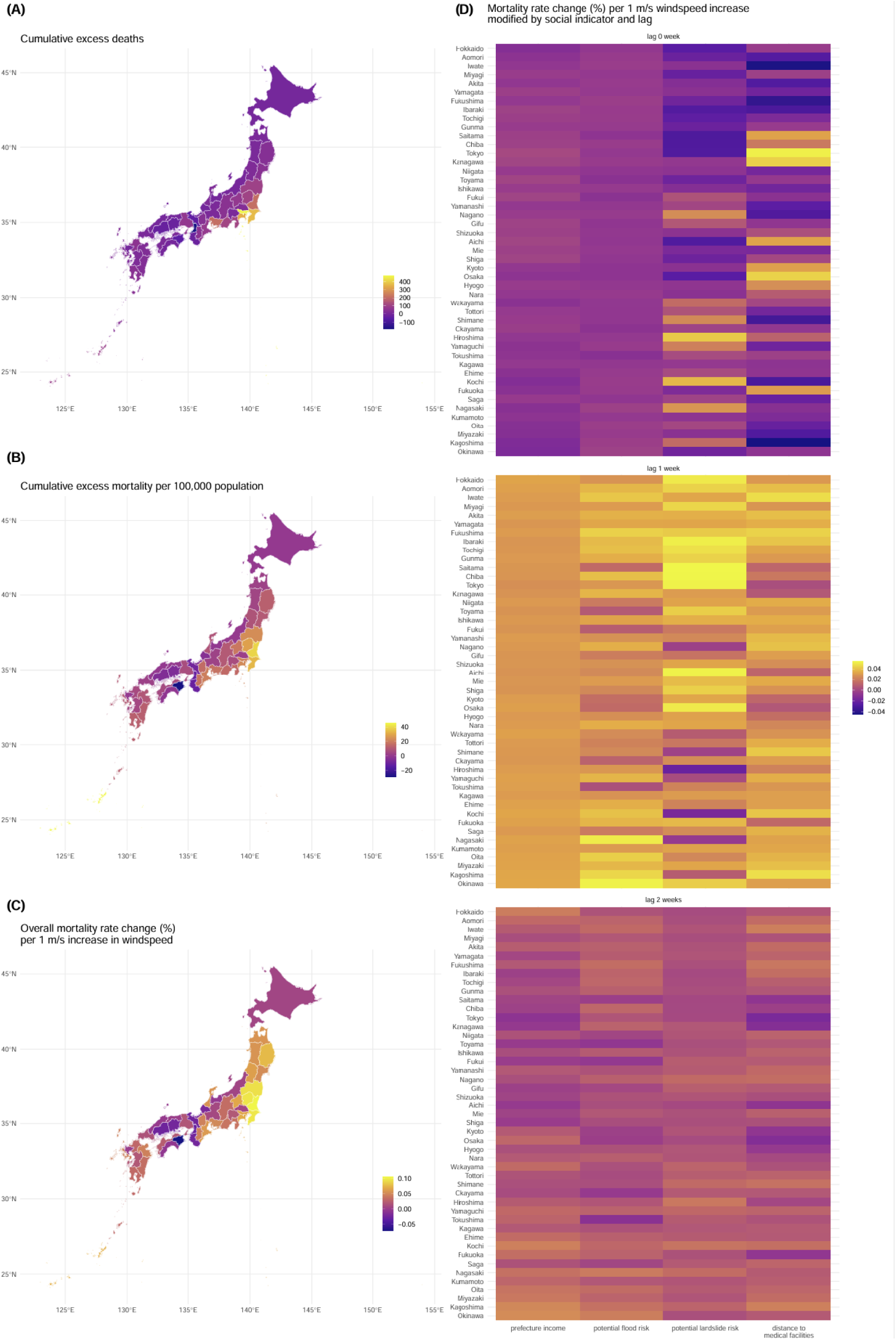
Typhoon-related mortality burden and its modification by socio-environmental factors in the population aged ≥70 years. (A) Cumulative excess deaths by prefecture, 2010–2019. (B) Cumulative excess mortality rate (%). (C) Overall mortality rate change (%) per 1 m/s increase in windspeed. (D) Modification of mortality risk (percent change per 1 m/s windspeed increase) by prefecture-level vulnerability indicators (income, flood risk, landslide risk, access to medical facilities) at 0, 1 and 2-week lags

Among adults ≥70 years, modifier effects on excess mortality were observed in the week of typhoon exposure for healthcare access and one week later for flood and landslide risk (Figure 4D), in contrast to the delayed two-week pattern of landslide and income effects observed among people <70 years. Socioeconomic and geographical factors exhibited stronger associations approximately one week after typhoon exposure (Figure 4D), in contrast to the younger population, in which a delayed response was detected, peaking at a two-week lag. In the week of typhoon exposure, healthcare access was the primary modifier across all prefectures, with the strongest association observed in major metropolitan areas. Tokyo showed the highest risk increase at 0.05% [95% CrI: 0.01, 0.10] per 1 m/s increase in windspeed (Figure 4D). One week after typhoon exposure, potential flood risk and landslide risk emerged as modifiers, with the greatest effects in southern and island prefectures; Okinawa showed the highest flood-related increase at 0.05% [95% CrI: 0.01, 0.09], and Chiba showed the largest landslide-related association at 0.05% [95% CrI: 0.02, 0.09]. Income was generally a weak modifier across all lags. Nagasaki and Okinawa showed the largest flood-related increases among older adults, with mortality increases of 0.05% [95% CrI: 0.01, 0.09] each.

## Discussion

This study estimated the nationwide, age-stratified typhoon-related excess mortality in Japan, a country combining one of the world’s highest tropical cyclone exposures with the most rapidly ageing population and quantified the nuanced role of underlying sociodemographic and geographical vulnerability factors in amplifying and/or attenuating the excess deaths in each lagged week associated with typhoons. The overall results suggest that those factors appeared to contribute to the increase in mortality observed among older adults during the first week of and the week following the typhoon exposure, while the effects were more pronounced in the second week for the younger group. It is plausible that their greater frailty and higher prevalence of chronic conditions result in more rapid typhoon-triggered excess deaths compared with younger individuals.

Among adults ≥70 years, excess mortality was concentrated in the immediate week of and the week following typhoon exposure, with flood risk and healthcare access as the primary modifiers, the latter showing higher excess mortality in metropolitan prefectures where baseline access was better. Prior studies have documented short-term post-cyclone mortality: in East Asia, respiratory mortality was highest on the day of typhoon exposure and declined to baseline within several days.^16^ In US adults aged ≥65 years, the largest mortality increases for injuries and several other causes occurred one month after exposure.^2^ Our weekly analysis identifies socio-geographic modifiers acting at lag 0–1 weeks, likely reflecting the greater physiological frailty of older adults and the higher burden of chronic conditions^17^ requiring continuous management, which renders this group particularly susceptible to short-term interruptions in healthcare access and to the immediate physiological stress imposed by severe weather events.

As opposed to the older group, among people <70 years, the modifier-attributable effects were delayed, peaking at two weeks, and were driven primarily by landslide risk and lower income. A previous multi-country analysis reported that relative increases in respiratory and neuropsychiatric mortality during the first two weeks after typhoon exposure were greater among individuals aged <60 years than among those aged ≥60 years, although the absolute burden of these deaths remains concentrated in older adults.^18^ The two-week peak observed in the present study is consistent with indirect mortality pathways that require time to manifest, including deferred care-seeking under occupational and economic constraints and progressive deterioration from initially non-fatal injuries. Our findings also highlight that these indirect pathways may be most pronounced in this age group with landslide risk and household income serving as the key modifiers.

In both younger and older age groups, shorter distances to medical facilities, indicating better medical access and typically observed in large metropolitan areas such as Tokyo and Osaka, were paradoxically associated with higher excess mortality during the week of typhoon exposure. Although our analysis used the proportion of households located more than 1,000 m from the nearest medical facility as a proxy indicator, this variable likely captures broader aspects of healthcare accessibility, including emergency transport time and the overall capacity to reach timely medical care. According to the national statistics, the average emergency transport time is 17.0 minutes in Tokyo and 24.2 minutes in Osaka, whereas it exceeds 90 minutes in Kagoshima and Wakayama,^19^ approximately a fivefold difference between the fastest and slowest prefectures. Moreover, heavy rainfall accompanying typhoons can itself delay emergency medical response; ambulance response times in Kyushu increased substantially once hourly rainfall exceeded approximately 30 mm, with severe cases being particularly sensitive to even moderate rainfall intensity.^20^ The stronger impact of typhoon exposure observed in prefectures with better medical access may therefore arise from the temporary loss of healthcare services that are usually readily available in these regions. Although hurricanes have been shown to increase emergency department visits,^21^ healthcare-seeking for minor conditions may be suppressed during the event,^22^ potentially contributing to fatal outcomes about one week later. When access to medical services is disrupted, conditions that are typically treatable may become fatal. This transient collapse of healthcare accessibility in otherwise well-resourced regions may paradoxically result in higher excess mortality compared with areas where baseline access is chronically limited. Such dynamics underscore the dual importance of maintaining surge capacity and ensuring continuity of care during extreme weather events.

A further finding with implications for disaster risk reduction is that typhoon-related excess mortality was not confined to coastal areas. The inland prefectures of Tochigi and Gunma ranked among the highest in cumulative excess deaths nationally among adults ≥70 years. As both prefectures ranked in the lower half nationally for typhoon-exposure frequency (10 weeks each), this burden likely reflects the extreme intensity of specific events—particularly Vongfong (2014) — combined with landslide risk, which was reflected in elevated landslide-related mortality at one week after typhoon exposure, as well as lack of previous direct experience in handling typhoon-related health risk. Given that approximately three-fourths of Japan’s land area is mountainous,^23^ inland areas are potentially highly susceptible to landslide-related mortality during typhoons. Disaster preparedness frameworks that focus primarily on coastal storm surge and wind damage may therefore underestimate mortality risk in mountainous inland areas.

Our findings on age-specific and geographically mediated risks are consistent with national reports from recent typhoon disasters in Japan. For example, official statistics from the Ministry of Land, Infrastructure, Transport and Tourism documented 82 deaths during Typhoon Talas (2011)^24^ and 96 during Typhoon Hagibis (2019);^25^ these figures reflect direct disaster deaths, yet a large proportion were attributable to landslides and floods in mountainous and riverine areas. These reports are broadly consistent with the patterns we identify. In both events, increases in deaths were observed not only at landfall itself but also approximately one week later. This temporal pattern suggests that mortality after typhoons reflects more than immediate trauma; it likely also includes exacerbations of chronic conditions among older evacuees, stress and infections associated with shelter living, and deaths occurring in previously isolated or unaccounted-for households. These mechanisms are also described in official disaster assessment documents, consistent with our findings. In addition, delays in discovering bodies and completing formal death certification may further contribute to the apparent lag.

The international Emergency Events Database (EM-DAT)^26,27^ and national disaster registries primarily capture direct deaths and may therefore substantially underestimate the broader mortality burden associated with typhoons. Over the 10-year study period, our model estimated a nationwide cumulative total of 2,426 excess deaths [139, 4,632] among adults aged ≥70 years, compared with 301 direct typhoon-related deaths recorded in EM-DAT over the same period (Supplementary Table 3). This discrepancy was also apparent across individual events. During Typhoon Vongfong in 2014, for example, EM-DAT recorded nine deaths nationwide, whereas the posterior median excess deaths reached 35 [95% CrI: 6, 64] in Chiba prefecture alone, and 148 deaths nationwide [95% CrI: −116, 449]. Although national-level credible intervals for single events often included zero, posterior median estimates generally exceeded direct death counts. These differences could be explained by a substantial contribution from indirect and delayed pathways, such as exacerbation of chronic conditions, evacuation-related stress, and disrupted continuity of care, which remain uncaptured by conventional disaster registries.

Although prefectural income disparities in Japan are relatively narrow compared with many other countries, our findings revealed that income still modified typhoon-related mortality risk among the younger population, specifically at a 2-week lag. This delayed pattern may reflect occupational and economic pressures faced by working-age adults in lower-income prefectures, where individuals may return more quickly to physically demanding outdoor work, or may delay seeking medical care^28^ due to limited workplace flexibility. In contrast, the absence of income-related effect modification on typhoon mortality among older adults may be partly attributable to Japan’s universal social protection systems,^29^ including national health insurance, the long-term care insurance programme and the medical care system for the latter-stage older population, which collectively buffer the influence of household income on healthcare access regardless of prefectural economic conditions.

There are several limitations to be noted. First, the model assumes that the reporting system and population structure remained constant over time, which may not fully capture changes in surveillance capacity or demographic composition during the study period. Second, although spatially structured and unstructured random effects were incorporated to account for regional heterogeneity, unmeasured local factors, such as differences in medical resource allocation, disaster preparedness, population density, or housing quality, may still bias the estimates. Third, the exposure metric was based on prefecture-level mean windspeed, which does not fully reflect within-prefecture variation in storm intensity or flooding. A finer spatial resolution of exposure data could help reduce potential misclassification. Fourth, while lag structures were explicitly modelled to represent delayed effects, the choice of maximum lag and the assumption of linearity in the windspeed–mortality association may have simplified complex temporal dynamics. Fifth, the age stratification used in this analysis was relatively coarse, as it classified individuals simply as below or above 70 years of age due to the limitation of the dataset, which may have masked important variations in vulnerability within narrower age groups. Finally, the model was designed to identify associations rather than causality; therefore, further causal analyses or mechanistic models integrating meteorological, infrastructural, and health system factors would be necessary to better elucidate the pathways linking typhoon exposure and mortality. Additionally, part of the excess mortality among older adults may reflect a mortality displacement (harvesting) effect, whereby typhoon exposure advances the timing of deaths among frail individuals. Such displacement would also be expected to manifest as a transient decrease in relative risk in subsequent weeks; further analyses discriminating harvesting from sustained risk would strengthen causal interpretation.

Taken together, our findings suggest that excess mortality linked to typhoons in high-income settings is shaped by the intersection of age-related frailty, disruptions in access to care and geographical factors. While hazard mapping, evacuation drills and infrastructure improvements are essential for preventing deaths immediately after a typhoon, our findings highlight the need for age-specific interventions. For younger adults, delayed vulnerability at two weeks was driven by landslide risk and lower income. Older adults experienced greater excess mortality, with vulnerability at lag 0–1 weeks driven by disruption of healthcare access, most evident in metropolitan prefectures where baseline access was better, and by flood and landslide risk. Consistent with these findings, practical responses such as expanding post-event access to care, including surge capacity in emergency services and support for medication management, may yield substantial benefits in averting excess mortality in vulnerable populations. Given the rapid population ageing in Japan, these measures are both feasible and immediately relevant for protecting older populations.

## Methods

### Study area and mortality data

We analysed data from all 47 prefectures in Japan during 2010–2019. Analyses were stratified by age (≥70 years vs. <70 years). We used weekly all-cause mortality, obtained from the Ministry of Health, Labour and Welfare of Japan. For comparison with our model-based estimates, we also compiled the number of typhoon-related deaths reported for Japan in EM-DAT, the international disaster database maintained by the Centre for Research on the Epidemiology of Disasters (CRED), UCLouvain^26,27^ (Supplementary Table 3).

### Exposure assessment

We quantified typhoon exposure using population-weighted sustained wind speed calculated from CLIMADA (CLIMate ADAptation),^30^ an open-source Python package. Typhoon tracks, maximum sustained wind speeds, and minimum central pressures were downloaded from the International Best Track Archive for Climate Stewardship, and the parametric windfield model of choice was Holland (1980).^4^ This metric integrates both wind intensity and its spatial extent, capturing variations in exposure between densely populated and sparsely populated areas, and recognising that wind-related damage can occur far from the storm centre.

For each prefecture, we defined typhoon exposure as any day when the maximum sustained wind speed at the population centroid reached 17.2 m/s (34 knots) or higher, based on Japan’s threshold for a tropical storm^31^. Here, we use the term typhoon exposure in a broader definition, including tropical storms, severe tropical storms, and typhoons, encompassing all storms that meet or exceed this wind threshold. A prefecture was classified as exposed for a given week if at least one day met this criterion; days below the threshold were treated as non-exposed.

### Covariates

#### Healthcare access

Limited spatial access to medical care can delay emergency response and continuity of care during typhoon-related disruptions. Access to medical care was represented by the proportion of households located more than 1,000 m from the nearest medical facility, as obtained from the Statistics Bureau of Japan (2018).^32^ This indicator serves as a proxy for limited healthcare accessibility and potential barriers to timely care during extreme weather events.

#### Geographical hazard exposure

To capture potential physical pathways linking typhoons to health risks, we used calculated proportions of residents living in flood- and landslide-prone areas from the Ministry of Land, Infrastructure, Transport and Tourism (MLIT), which was published for the disaster risk map in Japan.^33^ Briefly, flood-prone regions came from MLIT’s Inundation Assumption Areas (>0 m modelled inundation under design-scale rainfall), and landslide-prone regions from MLIT’s Sediment Disaster Hazard Areas (designated debris-flow/steep-slope zones). Residents in at-risk cells were then summed and expressed as a proportion of the total prefectural population.

#### Income

Prefecture-level income data were collected from the 2019 National Survey of Family Income and Expenditure (Statistics Bureau of Japan).^34^ Household income captures adaptive capacity (e.g., resilient housing). We used the mean annual household income for each prefecture, irrespective of household size, worker/ordinary household classification, or sex.

## Statistical analysis

We reconstructed the process of generating new deaths per prefecture with and without typhoon exposures in week t to estimate the contribution of underlying factors on the typhoon-associated excess deaths. We assumed the weekly number of deaths at prefecture p, *Y_n_*_,*p*_, follows a Poisson distribution as below:

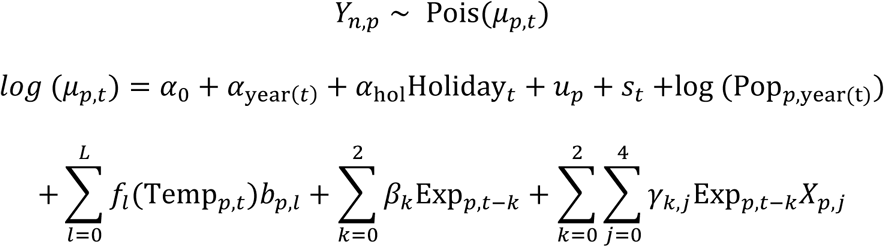

This model can be partitioned into two parts: the first bulk representing a baseline scenario of deaths in which there is no typhoon exposure, and the second component that explains the typhoon-associated deaths accounting for delayed effects of the exposure and several potential driving factors.

To predict the baseline number of deaths, several key measurements are included in the model, such as the global and year-specific intercept, *α*_0_ and *α*_year(*t*)_. We also incorporated the impact of holidays, Holiday*_t_*, with the associated coefficient, *α*_hol_, to capture the short-term disruptions in mortality patterns. As the analysis included all 47 prefectures, accounting for geographical variation is also important given the potential correlation in mortality risk across neighbouring regions. To address this, we incorporated spatially structured random effects following an intrinsic conditional autoregressive (ICAR) model on the variable, *u_p_*. A weekly temporal random effect, *s_t_*, is included, modelled with a second-order random walk prior to track seasonal variation in mortality risk. In addition, given that temperature is a major predictor of mortality,^35^ we modelled the nonlinear effects of weekly mean temperature, Temp*_p_*_,*t*_, specifying prefecture-specific cubic B-spline functions, *f_l_*(⋅), with its associated linear coefficient, *b_p_*_,*l*_. Finally, the term log (Pop*_p_*_,year(t)_) is the model offset with the yearly population size at prefecture, p.

Building upon the aforementioned baseline component of the model, the remaining part captures the excess mortality attributable to typhoon exposure. To account for the temporal propagation of wind effects, we employed the convolution structure that combines both immediate and delayed effects across successive two weeks (0-2 weeks): *Σ^2^_k=0_β_k_Exp_p,t−k_*, where Exp*_p_*_,*t*_denotes the typhoon exposures defined as the estimated temporal maximum windspeed (m/s) within each prefecture, and *β_k_* its associated coefficient. To capture how local conditions amplify or attenuate the mortality attributable to typhoon exposure, we included the interactions between lagged exposure and prefecture-specific sociodemographic and environmental indicators: *Σ^2^_k=0_Σ^4^_j=0_γ_k_Exp_p,t−k_X_p,j_*, where *X_p_*_,*j*_ represents four standardised time-invariant indicators (j=1: income, 2: flood-prone area percentage, 3: landslide-prone area percentage and 4: proportion of households located more than 1000 meters from the nearest medical facility).

To quantify how mortality changes with increasing windspeed, we computed the percent change in mortality associated with a 1 m/s increase in windspeed for each prefecture. This is derived by exponentiating the combined lag-specific effects and subtracting one:

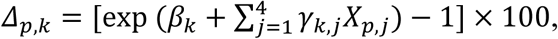

where *Δ* represents the percentage change in mortality corresponding to windspeed at lag k.

The overall percentage change across all the lags was then obtained by aggregating over k:

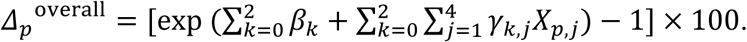

Parameter estimation was conducted within a Bayesian framework using the integrated nested Laplace approximation. This approach provides a computationally efficient analytical posterior approximation, making it well-suited for high-dimensional spatio-temporal models and allowing robust inference. We assigned Gaussian priors Normal(0, 1000) to fixed effect coefficients, with a flat prior for the intercept. Precision parameters for latent random effects were assigned log-Gamma priors on the log-precision scale with shape 1 and 5×10^-5^. To support interpretation under uncertainty, we report both 95% credible intervals and the posterior probability that excess deaths were greater than zero.

## Code availability

All the analysis was conducted in R v.4.5.0. Source codes with a dummy dataset are available on a GitHub repository:https://github.com/hiroaki-murayama/typhoon_social_jp.

## Author contributions

LY and HM conceived the study. LY and PLC curated datasets. LY, HM, MH, and RMP contributed to the study design. LY and HM conducted data analysis and drafted the original manuscript. RMP supervised the study. All authors contributed to reviewing and editing the manuscript and approved the final version.

## Data Availability

Individual-level mortality data are not publicly available due to privacy protection mandates under the Statistics Act of Japan. These data were provided by the Ministry of Health, Labour and Welfare (MHLW) under a specific license for this study. Qualified researchers may request access to these data directly from the MHLW at https://www.mhlw.go.jp/toukei_jouhou/use/index.html. Meteorological data regarding temperature and humidity can be downloaded from the Japan Meteorological Agency (JMA) website at https://www.data.jma.go.jp/stats/etrn/index.php. Tropical cyclone wind speed data were retrieved from the International Best Track Archive for Climate Stewardship (IBTrACS) at https://www.ncei.noaa.gov/products/international-best-track-archive. Data on the distance to medical facilities were derived from the Housing and Land Survey, available via the Portal Site of Official Statistics of Japan (e-Stat) at https://www.e-stat.go.jp/stat-search/files?page=1&toukei=00200522. Estimates of landslide and flood-prone areas were obtained from the Ministry of Land, Infrastructure, Transport and Tourism (MLIT) at https://www.mlit.go.jp/kokudoseisaku/content/001373119.pdf.

## Acknowledgements

LY is supported by the Fulbright Foreign Student Program and by JSPS KAKENHI Grant Number JP24K13497. Robbie M Parks is supported by Wellcome Trust grants 327930/Z/25/Z, 324337/Z/25/Z, 335293/Z/25/Z, and NIH grants R00 ES033742, D43 TW012726, and NFRF grant NFRFI-2023-00222. This research was performed by the Environment Research and Technology Development Fund (JPMEERF25S12400 and JPMEERF20262001) of the Environmental Restoration and Conservation Agency provided by Ministry of the Environment of Japan.

