## Supplementary Information for "Socio-demographic and environmental factors amplify typhoon-related excess mortality in Japan"

Lisa Yamasaki[1,2,3‡], Hiroaki Murayama[4‡], Paul LC Chua[3], Masahiro Hashizume[3,5], Robbie M. Parks[6*]

[1]Department of Environmental Health, Harvard T.H. Chan School of Public Health, Boston, MA, USA

[2]Department of Emergency Medicine and Intensive Care Unit, Center Hospital of the National Center for Global Health and Medicine, Tokyo, Japan

[3]Department of Global Health Policy, Graduate School of Medicine, The University of Tokyo, Tokyo, Japan

[4] School of Medicine, International University of Health and Welfare, Narita, Japan

[5]School of Tropical Medicine and Global Health, Nagasaki University, Nagasaki, Japan

[6]Department of Environmental Health Sciences, Mailman School of Public Health, Columbia University, New York, USA

‡ These authors equally contributed to this work

**Supplementary Material**

Socio-demographic and environmental factors amplify typhoon-related excess mortality in Japan

### Supplementary Tables

- Supplementary Table 1. Prefecture- and typhoon-specific excess deaths with posterior probability >95% for population aged <70 years
- Supplementary Table 2. Prefecture- and typhoon-specific excess deaths with posterior probability >95% for population aged ≥70 years
- Supplementary Table 3. National excess deaths by typhoon, 2010–2019.

### Supplementary Figures

- Supplementary Figure 1. Socioeconomic and geographical vulnerability indicators by prefecture (Income, landslide-prone areas, flood-prone areas, access to medical facilities)
- Supplementary Figure 2. Posterior probability that excess deaths exceed zero by age group

**Supplementary Table 1.** Prefecture- and typhoon-specific excess deaths with posterior probability >95% for population aged <70 years, 2010–2019.

| Prefecture | Year | Typhoon | Estimated excess deaths | Posterior probability* |
|---|---|---|---|---|
| Aomori | 2016 | Namtheun | 2 [0, 3] | 97.8% |
| Aomori | 2018 | Jebi | 1 [0, 2] | 97.8% |
| Iwate | 2010 | Dianmu | 1 [0, 2] | 99.3% |
| Iwate | 2012 | Jelawat | 1 [0, 3] | 99.3% |
| Iwate | 2013 | Man-yi | 1 [0, 2] | 99.3% |
| Iwate | 2014 | Vongfong | 1 [0, 2] | 99.3% |
| Iwate | 2016 | Mindulle | 1 [0, 2] | 99.3% |
| Iwate | 2016 | Namtheun | 2 [1, 4] | 100.0% |
| Iwate | 2018 | Trami | 1 [0, 2] | 99.3% |
| Iwate | 2019 | Hagibis | 1 [0, 2] | 99.3% |
| Miyagi | 2010 | Dianmu | 1 [0, 2] | 96.5% |
| Miyagi | 2011 | Roke | 1 [0, 2] | 96.5% |
| Miyagi | 2012 | Jelawat | 2 [0, 4] | 96.5% |
| Miyagi | 2013 | Man-yi | 1 [0, 2] | 96.5% |
| Miyagi | 2014 | Vongfong | 1 [0, 2] | 96.5% |
| Miyagi | 2016 | Mindulle | 2 [0, 4] | 96.3% |
| Miyagi | 2016 | Namtheun | 2 [0, 4] | 98.6% |
| Miyagi | 2018 | Trami | 1 [0, 2] | 96.5% |
| Miyagi | 2019 | Hagibis | 1 [0, 2] | 96.5% |
| Akita | 2010 | Dianmu | 1 [0, 1] | 98.9% |
| Akita | 2016 | Namtheun | 1 [0, 1] | 98.9% |
| Akita | 2018 | Jebi | 1 [0, 1] | 98.9% |
| Akita | 2019 | Hagibis | 0 [0, 1] | 98.9% |
| Yamagata | 2010 | Dianmu | 1 [0, 1] | 99.5% |
| Yamagata | 2012 | Jelawat | 1 [0, 2] | 99.5% |
| Yamagata | 2013 | Man-yi | 0 [0, 1] | 99.5% |
| Yamagata | 2014 | Vongfong | 0 [0, 1] | 99.5% |
| Yamagata | 2018 | Jebi | 0 [0, 1] | 99.5% |
| Yamagata | 2018 | Trami | 0 [0, 1] | 99.5% |
| Yamagata | 2019 | Hagibis | 1 [0, 1] | 99.5% |
| Fukushima | 2011 | Roke | 2 [1, 4] | 99.8% |
| Fukushima | 2012 | Guchol | 2 [1, 4] | 99.8% |
| Fukushima | 2012 | Jelawat | 4 [1, 6] | 99.8% |
| Fukushima | 2013 | Man-yi | 3 [1, 4] | 99.8% |
| Fukushima | 2014 | Vongfong | 2 [1, 3] | 99.8% |
| Fukushima | 2016 | Mindulle | 4 [1, 6] | 100.0% |
| Fukushima | 2017 | Lan | 2 [1, 3] | 99.8% |
| Fukushima | 2018 | Leepi | 1 [0, 2] | 99.8% |
| Fukushima | 2018 | Trami | 3 [1, 4] | 99.8% |
| Fukushima | 2019 | Hagibis | 2 [1, 4] | 99.8% |
| Ibaraki | 2011 | Roke | 6 [1, 11] | 99.8% |
| Ibaraki | 2012 | Guchol | 4 [1, 8] | 99.8% |
| Ibaraki | 2012 | Jelawat | 4 [1, 7] | 99.8% |

| | | | | |
|---|---|---|---|---|
| Ibaraki | 2013 | Man-yi | 4 [1, 8] | 99.8% |
| Ibaraki | 2013 | Wipha | 3 [1, 6] | 99.8% |
| Ibaraki | 2014 | Vongfong | 10 [2, 18] | 99.1% |
| Ibaraki | 2016 | Mindulle | 7 [2, 11] | 99.4% |
| Ibaraki | 2017 | Lan | 5 [1, 9] | 99.8% |
| Ibaraki | 2018 | Leepi | 2 [0, 4] | 99.8% |
| Ibaraki | 2018 | Trami | 3 [1, 6] | 99.8% |
| Ibaraki | 2019 | Faxai | 3 [1, 5] | 99.8% |
| Ibaraki | 2019 | Hagibis | 4 [1, 8] | 99.8% |
| Tochigi | 2011 | Roke | 3 [1, 5] | 99.9% |
| Tochigi | 2012 | Guchol | 3 [1, 5] | 99.9% |
| Tochigi | 2012 | Jelawat | 3 [1, 6] | 99.9% |
| Tochigi | 2013 | Man-yi | 3 [1, 5] | 99.9% |
| Tochigi | 2013 | Wipha | 2 [0, 2] | 99.9% |
| Tochigi | 2014 | Vongfong | 4 [1, 8] | 99.3% |
| Tochigi | 2016 | Mindulle | 2 [0, 3] | 99.9% |
| Tochigi | 2017 | Lan | 3 [1, 5] | 99.9% |
| Tochigi | 2018 | Trami | 3 [1, 5] | 99.9% |
| Tochigi | 2019 | Hagibis | 2 [1, 4] | 99.9% |
| Gunma | 2011 | Roke | 2 [0, 3] | 99.6% |
| Gunma | 2012 | Guchol | 2 [0, 3] | 99.6% |
| Gunma | 2012 | Jelawat | 2 [0, 4] | 99.6% |
| Gunma | 2013 | Man-yi | 2 [0, 3] | 99.6% |
| Gunma | 2014 | Vongfong | 3 [0, 6] | 98.6% |
| Gunma | 2016 | Mindulle | 1 [0, 2] | 99.6% |
| Gunma | 2017 | Lan | 2 [0, 3] | 99.6% |
| Gunma | 2017 | Talim | 1 [0, 2] | 99.6% |
| Gunma | 2018 | Trami | 2 [0, 4] | 99.6% |
| Gunma | 2019 | Hagibis | 1 [0, 2] | 99.6% |
| Chiba | 2011 | Roke | 10 [0, 19] | 98.4% |
| Chiba | 2012 | Guchol | 6 [0, 11] | 98.4% |
| Chiba | 2012 | Jelawat | 6 [0, 11] | 98.4% |
| Chiba | 2013 | Man-yi | 6 [0, 12] | 98.4% |
| Chiba | 2013 | Wipha | 6 [0, 11] | 98.4% |
| Chiba | 2017 | Lan | 11 [0, 22] | 98.4% |
| Chiba | 2018 | Leepi | 4 [0, 7] | 98.4% |
| Chiba | 2018 | Trami | 5 [0, 9] | 98.4% |
| Chiba | 2019 | Faxai | 6 [0, 12] | 98.4% |
| Chiba | 2019 | Hagibis | 9 [0, 18] | 98.4% |
| Ishikawa | 2011 | Roke | 0 [0, 1] | 99.0% |
| Ishikawa | 2013 | Man-yi | 0 [0, 1] | 99.0% |
| Ishikawa | 2014 | Halong | 1 [0, 1] | 99.0% |
| Ishikawa | 2014 | Vongfong | 0 [0, 1] | 99.0% |
| Ishikawa | 2015 | Etau | 0 [0, 1] | 99.0% |
| Ishikawa | 2017 | Lan | 0 [0, 1] | 99.0% |
| Ishikawa | 2017 | Talim | 0 [0, 1] | 99.0% |
| Ishikawa | 2018 | Jebi | 1 [0, 2] | 99.0% |
| Yamanashi | 2014 | Vongfong | 1 [0, 3] | 97.9% |

| | | | | |
|---|---|---|---|---|
| Mie | 2011 | Roke | 1 [0, 2] | 98.1% |
| Mie | 2012 | Guchol | 1 [0, 2] | 98.1% |
| Mie | 2012 | Jelawat | 1 [0, 2] | 98.1% |
| Mie | 2013 | Man-yi | 1 [0, 2] | 98.1% |
| Mie | 2014 | Vongfong | 3 [0, 5] | 98.5% |
| Mie | 2016 | Malakas | 1 [0, 2] | 98.1% |
| Mie | 2017 | Lan | 1 [0, 2] | 98.1% |
| Mie | 2017 | Noru | 1 [0, 2] | 98.1% |
| Mie | 2017 | Talim | 1 [0, 3] | 98.1% |
| Mie | 2018 | Jebi | 1 [0, 2] | 98.1% |
| Mie | 2018 | Jongdari | 1 [0, 2] | 98.1% |
| Mie | 2018 | Trami | 1 [0, 3] | 98.1% |
| Nara | 2011 | Ma-on | 0 [0, 1] | 97.8% |
| Nara | 2011 | Roke | 1 [0, 2] | 98.4% |
| Nara | 2011 | Talas | 0 [0, 1] | 97.8% |
| Nara | 2012 | Guchol | 1 [0, 1] | 97.8% |
| Nara | 2012 | Jelawat | 1 [0, 1] | 97.8% |
| Nara | 2013 | Man-yi | 1 [0, 1] | 97.8% |
| Nara | 2014 | Halong | 0 [0, 1] | 97.8% |
| Nara | 2014 | Vongfong | 1 [0, 3] | 95.5% |
| Nara | 2016 | Malakas | 0 [0, 1] | 97.8% |
| Nara | 2017 | Lan | 0 [0, 1] | 97.8% |
| Nara | 2017 | Noru | 1 [0, 1] | 97.8% |
| Nara | 2017 | Talim | 1 [0, 2] | 97.8% |
| Nara | 2018 | Jebi | 2 [0, 4] | 96.1% |
| Nara | 2018 | Jongdari | 1 [0, 2] | 97.8% |
| Nara | 2018 | Rumbia | 0 [0, 1] | 97.8% |
| Nara | 2018 | Trami | 1 [0, 1] | 97.8% |
| Nara | 2019 | Krosa | 0 [0, 1] | 97.8% |
| Kagawa | 2015 | Halola | 1 [0, 2] | 95.5% |
| Kagawa | 2018 | Jebi | 1 [0, 1] | 97.1% |
| Saga | 2019 | Krosa | 0 [0, 1] | 98.6% |
| Kumamoto | 2018 | Rumbia | 1 [0, 2] | 99.3% |
| Kumamoto | 2019 | Krosa | 1 [0, 2] | 99.1% |
| Oita | 2011 | Ma-on | 1 [0, 1] | 95.6% |
| Oita | 2014 | Halong | 0 [0, 1] | 95.6% |
| Oita | 2014 | Vongfong | 1 [0, 2] | 95.6% |
| Oita | 2015 | Goni | 1 [0, 2] | 95.6% |
| Oita | 2016 | Malakas | 1 [0, 1] | 95.6% |
| Oita | 2017 | Nanmadol | 0 [0, 1] | 95.6% |
| Oita | 2017 | Talim | 1 [0, 1] | 95.6% |
| Oita | 2018 | Rumbia | 0 [0, 1] | 95.6% |
| Oita | 2018 | Trami | 1 [0, 2] | 95.6% |
| Oita | 2019 | Krosa | 1 [0, 1] | 95.6% |
| Oita | 2019 | Tapah | 0 [0, 1] | 95.6% |
| Miyazaki | 2014 | Vongfong | 2 [0, 4] | 97.8% |
| Miyazaki | 2019 | Krosa | 1 [0, 2] | 99.3% |
| Kagoshima | 2014 | Vongfong | 4 [-1, 8] | 95.1% |

| | | | | |
|---|---|---|---|---|
| Kagoshima | 2016 | Malakas | 2 [0, 5] | 95.1% |
| Okinawa | 2010 | Malou | 4 [-1, 8] | 96.0% |
| Okinawa | 2011 | Aere | 1 [0, 3] | 95.9% |
| Okinawa | 2011 | Muifa | 4 [0, 9] | 96.0% |
| Okinawa | 2011 | Songda | 3 [0, 6] | 95.9% |
| Okinawa | 2012 | Bolaven | 2 [0, 5] | 95.9% |
| Okinawa | 2012 | Guchol | 1 [0, 3] | 95.9% |
| Okinawa | 2012 | Jelawat | 10 [2, 18] | 99.4% |
| Okinawa | 2013 | Danas | 1 [0, 3] | 95.9% |
| Okinawa | 2013 | Francisco | 3 [1, 4] | 99.6% |
| Okinawa | 2014 | Nakri | 3 [0, 7] | 96.0% |
| Okinawa | 2014 | Neoguri | 1 [0, 2] | 95.9% |
| Okinawa | 2014 | Vongfong | 2 [0, 5] | 95.9% |
| Okinawa | 2015 | Goni | 1 [0, 3] | 95.9% |
| Okinawa | 2015 | Halola | 1 [0, 2] | 95.9% |
| Okinawa | 2016 | Chaba | 2 [0, 3] | 95.9% |
| Okinawa | 2017 | Saola | 2 [0, 4] | 95.9% |
| Okinawa | 2017 | Talim | 1 [0, 2] | 95.9% |
| Okinawa | 2018 | Gaemi | 1 [0, 3] | 95.9% |
| Okinawa | 2018 | Jongdari | 2 [1, 4] | 99.9% |
| Okinawa | 2018 | Prapiroon | 3 [1, 5] | 99.9% |
| Okinawa | 2018 | Trami | 5 [-1, 10] | 95.9% |
| Okinawa | 2019 | Tapah | 2 [0, 4] | 95.9% |

*CrI:credible interval*

Posterior median and 95% credible interval (CrI) for cumulative excess deaths over a 0–2 week lag following typhoon exposure. Estimates with posterior probability that excess deaths exceeded zero >95% are shown.

**Supplementary Table 2.** Prefecture- and typhoon-specific excess deaths with posterior probability >95% for population aged ≥70 years, 2010–2019.

| Prefecture | Year | Typhoon | Estimated excess deaths | Posterior probability* |
|---|---|---|---|---|
| Miyagi | 2016 | Namtheun | 3 [0, 7] | 97.5% |
| Fukushima | 2016 | Mindulle | 5 [0, 11] | 96.8% |
| Ibaraki | 2014 | Vongfong | 13 [-2, 26] | 96.3% |
| Ibaraki | 2016 | Mindulle | 9 [-1, 18] | 96.3% |
| Tochigi | 2011 | Roke | 3 [0, 6] | 97.1% |
| Tochigi | 2012 | Guchol | 3 [0, 6] | 97.1% |
| Tochigi | 2012 | Jelawat | 4 [0, 7] | 97.1% |
| Tochigi | 2013 | Man-yi | 4 [0, 7] | 97.1% |
| Tochigi | 2013 | Wipha | 2 [0, 4] | 97.1% |
| Tochigi | 2014 | Vongfong | 7 [1, 13] | 98.4% |
| Tochigi | 2016 | Mindulle | 2 [0, 4] | 97.1% |
| Tochigi | 2017 | Lan | 4 [0, 8] | 97.1% |
| Tochigi | 2018 | Trami | 4 [0, 9] | 97.1% |
| Tochigi | 2019 | Hagibis | 4 [0, 8] | 97.1% |
| Gunma | 2011 | Roke | 2 [0, 4] | 95.4% |
| Gunma | 2012 | Guchol | 2 [0, 4] | 95.4% |
| Gunma | 2012 | Jelawat | 3 [-1, 6] | 95.4% |
| Gunma | 2013 | Man-yi | 2 [0, 5] | 95.4% |
| Gunma | 2014 | Vongfong | 5 [0, 10] | 97.8% |
| Gunma | 2016 | Mindulle | 1 [0, 3] | 95.4% |
| Gunma | 2017 | Lan | 3 [-1, 7] | 95.4% |
| Gunma | 2017 | Talim | 2 [0, 3] | 95.4% |
| Gunma | 2018 | Trami | 3 [-1, 7] | 95.4% |
| Gunma | 2019 | Hagibis | 2 [0, 5] | 95.4% |
| Chiba | 2011 | Roke | 15 [2, 28] | 99.1% |
| Chiba | 2012 | Guchol | 9 [1, 16] | 99.1% |
| Chiba | 2012 | Jelawat | 10 [1, 17] | 99.1% |
| Chiba | 2013 | Man-yi | 12 [2, 21] | 99.1% |
| Chiba | 2013 | Wipha | 10 [2, 19] | 99.1% |
| Chiba | 2014 | Vongfong | 35 [6, 64] | 98.8% |
| Chiba | 2016 | Mindulle | 23 [4, 41] | 99.0% |
| Chiba | 2017 | Lan | 25 [4, 46] | 99.1% |
| Chiba | 2018 | Leepi | 9 [1, 16] | 99.1% |
| Chiba | 2018 | Trami | 11 [2, 20] | 99.1% |
| Chiba | 2019 | Faxai | 17 [2, 31] | 99.1% |
| Chiba | 2019 | Hagibis | 23 [3, 42] | 99.1% |
| Nara | 2018 | Jebi | 5 [0, 11] | 97.1% |
| Fukuoka | 2019 | Krosa | 7 [-1, 15] | 95.5% |
| Okinawa | 2010 | Malou | 5 [0, 10] | 97.0% |
| Okinawa | 2011 | Aere | 2 [0, 4] | 95.4% |
| Okinawa | 2011 | Muifa | 5 [0, 11] | 96.9% |
| Okinawa | 2011 | Songda | 3 [-1, 6] | 95.4% |
| Okinawa | 2012 | Bolaven | 3 [-1, 6] | 95.4% |

| | | | | |
|---|---|---|---|---|
| Okinawa | 2012 | Guchol | 2 [0, 3] | 95.4% |
| Okinawa | 2013 | Danas | 2 [0, 4] | 95.4% |
| Okinawa | 2014 | Nakri | 5 [0, 10] | 97.0% |
| Okinawa | 2014 | Neoguri | 2 [0, 3] | 95.4% |
| Okinawa | 2014 | Vongfong | 3 [-1, 6] | 95.4% |
| Okinawa | 2015 | Goni | 2 [0, 4] | 95.4% |
| Okinawa | 2015 | Halola | 2 [0, 3] | 95.4% |
| Okinawa | 2016 | Chaba | 2 [0, 5] | 95.4% |
| Okinawa | 2017 | Saola | 2 [0, 5] | 95.4% |
| Okinawa | 2017 | Talim | 2 [0, 3] | 95.4% |
| Okinawa | 2018 | Gaemi | 2 [0, 4] | 95.4% |
| Okinawa | 2018 | Maria | 3 [0, 6] | 96.3% |
| Okinawa | 2018 | Trami | 8 [0, 17] | 97.3% |
| Okinawa | 2019 | Tapah | 3 [-1, 6] | 95.4% |

*CrI:credible interval*

Posterior median and 95% credible interval (CrI) for cumulative excess deaths over a 0–2 week lag following typhoon exposure. Estimates with posterior probability that excess deaths exceeded zero >95% are shown.

**Supplementary Table 3.** National excess deaths by typhoon, 2010–2019.

| Year | Typhoon | Aged <70 years (Median [95% CrI]) | Aged ≥70 years (Median [95% CrI]) | EM-DAT deaths |
|---|---|---|---|---|
| 2010 | Dianmu | 4 [1, 7] | 1 [-5, 7] | – |
| 2010 | Malou | 4 [-1, 8] | 3 [-2, 8] | – |
| 2011 | Aere | 1 [0, 3] | 2 [0, 3] | – |
| 2011 | Ma-On | -1 [-15, 15] | -16 [-40, 10] | – |
| 2011 | Muifa | 4 [0, 9] | 5 [0, 11] | – |
| 2011 | Roke | 50 [-32, 139] | 52 [-74, 187] | 13 |
| 2011 | Songda | 3 [0, 6] | 3 [-1, 7] | – |
| 2011 | Talas | -1 [-12, 10] | -14 [-31, 6] | 68 |
| 2012 | Bolaven | 2 [0, 5] | 3 [-1, 6] | 0 |
| 2012 | Guchol | 35 [-26, 99] | 43 [-53, 140] | – |
| 2012 | Haikui | 1 [-1, 3] | -1 [-5, 3] | – |
| 2012 | Jelawat | 51 [-14, 121] | 50 [-54, 161] | 2 |
| 2013 | Danas | 4 [-5, 12] | -9 [-26, 8] | – |
| 2013 | Francisco | 3 [1, 5] | 1 [-2, 4] | – |
| 2013 | Man-Yi | 40 [-28, 109] | 47 [-72, 168] | 6 |
| 2013 | Toraji | 2 [-1, 6] | -2 [-9, 6] | 1 |
| 2013 | Wipha | 20 [-10, 53] | 37 [-13, 90] | 39 |
| 2014 | Halong | 0 [-16, 18] | -21 [-52, 14] | 10 |
| 2014 | Nakri | 3 [-1, 7] | 5 [0, 10] | 0 |
| 2014 | Neoguri | 4 [-1, 9] | 0 [-9, 10] | 7 |
| 2014 | Vongfong | 55 [-93, 224] | 148 [-116, 449] | 9 |
| 2015 | Etau | 4 [-4, 12] | -1 [-18, 15] | 8 |
| 2015 | Goni | 10 [-5, 25] | -4 [-36, 29] | 0 |
| 2015 | Halola | 5 [-3, 15] | -12 [-31, 9] | – |
| 2016 | Chaba | 2 [-2, 6] | -6 [-15, 3] | 0 |
| 2016 | Malakas | 7 [-11, 26] | -11 [-51, 29] | 1 |
| 2016 | Mindulle | 33 [-6, 73] | 69 [-3, 145] | 2 |
| 2016 | Namtheun | 7 [2, 12] | 7 [-4, 17] | – |
| 2017 | Lan | 49 [-33, 135] | 91 [-85, 275] | 8 |
| 2017 | Nanmadol | 2 [-1, 6] | -2 [-11, 7] | – |
| 2017 | Noru | 3 [-14, 20] | -12 [-47, 24] | 2 |
| 2017 | Saola | 2 [0, 4] | 2 [-1, 5] | – |
| 2017 | Talim | 13 [-22, 49] | -20 [-102, 63] | 2 |
| 2018 | Gaemi | 1 [0, 3] | 2 [0, 4] | – |
| 2018 | Jebi | -2 [-59, 56] | -21 [-154, 113] | 17 |
| 2018 | Jongdari | 8 [-18, 37] | -12 [-70, 50] | – |
| 2018 | Leepi | 7 [2, 12] | 13 [1, 25] | – |
| 2018 | Maria | 1 [-1, 3] | 3 [0, 6] | – |
| 2018 | Prapiroon | 4 [1, 7] | 1 [-3, 7] | – |
| 2018 | Rumbia | 0 [-14, 15] | -15 [-48, 22] | – |
| 2018 | Trami | 46 [-18, 114] | 53 [-89, 205] | 4 |
| 2019 | Faxai | 19 [-10, 51] | 53 [-15, 125] | 3 |
| 2019 | Francisco | 2 [-1, 6] | 2 [-7, 11] | – |

| | | | | |
|---|---|---|---|---|
| 2019 | Hagibis | 39 [-11, 92] | 82 [-36, 203] | 99 |
| 2019 | Krosa | 4 [-15, 23] | -20 [-70, 29] | – |
| 2019 | Nari | 2 [-3, 6] | 5 [-5, 15] | – |
| 2019 | Tapah | 7 [-5, 18] | -10 [-41, 21] | 0 |

Posterior median and 95% credible interval (CrI) of excess deaths obtained by aggregating prefecture-level posterior estimates across all 47 prefectures for each typhoon. All 47 typhoons that exceeded the 17.2 m/s population-weighted sustained wind speed exposure threshold during 2010–2019 are listed. Emergency Events Database (EM-DAT) direct deaths refer to nationwide fatalities reported by the Centre for Research on the Epidemiology of Disasters (CRED).

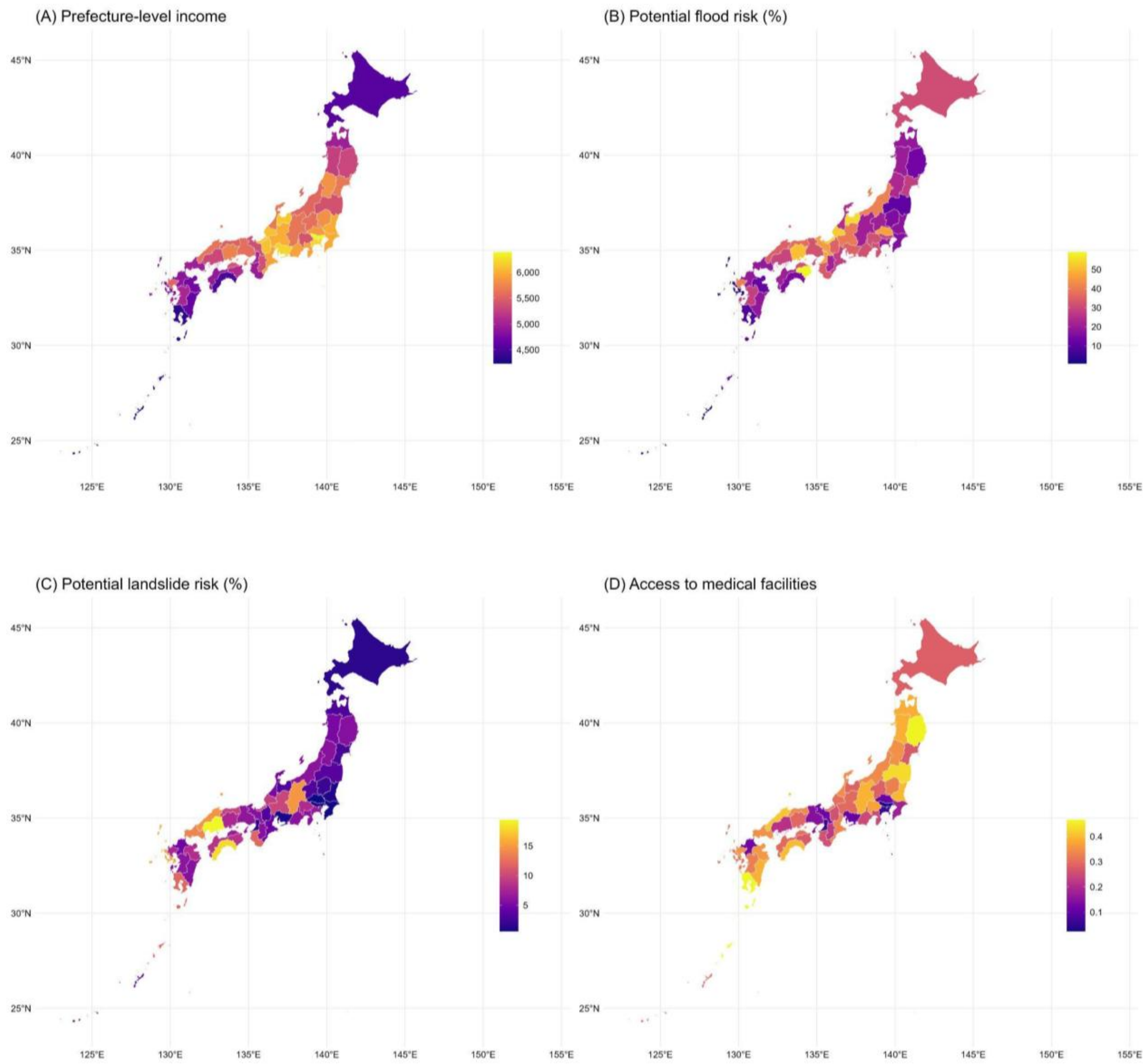


**Supplementary Figure 1.** Socioeconomic and geographical vulnerability indicators by prefecture (Income, landslide-prone areas, flood-prone areas, access to medical facilities). This figure illustrates the spatial distribution of four vulnerability metrics across Japan's 47 prefectures. Panel (A) shows per capita income, reflecting socioeconomic status. Panels (B) and (C) present the percentage of the population residing in flood-prone and landslide-prone areas, respectively, as of 2015. Panel (D) depicts an index of access to medical facilities.

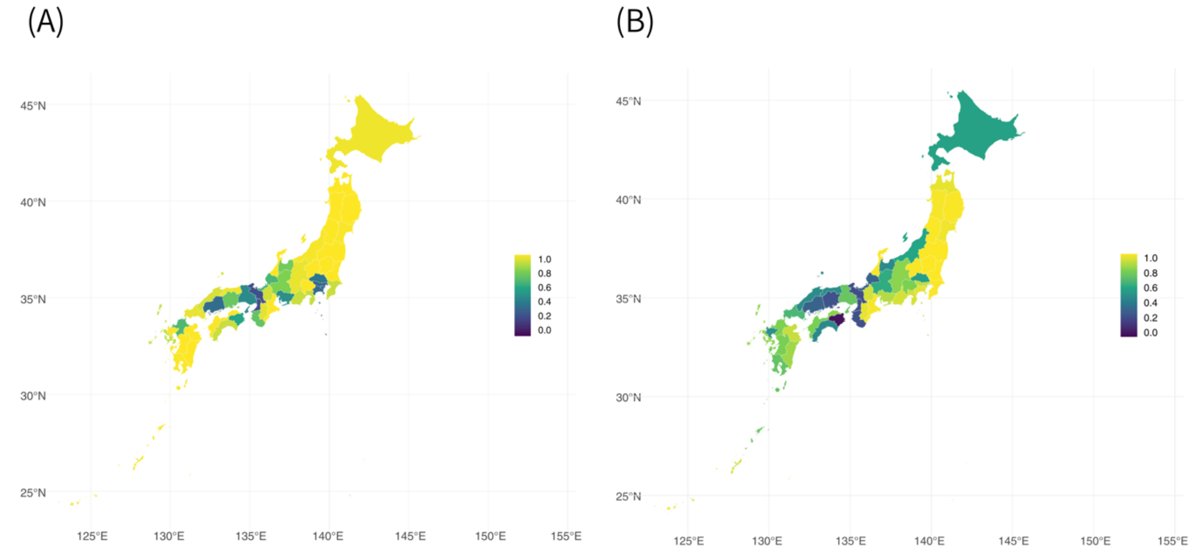


**Supplementary Figure 2.** Posterior probability that excess deaths exceed zero by age group. The posterior probability that excess deaths exceed zero is shown for individuals aged <70 years (A) and ≥70 years (B).
